# Demographic and co-morbidity characteristics of patients tested for SARS-CoV-2 from March 2020 to January 2022 in a national clinical research network: results from PCORnet^®^

**DOI:** 10.1101/2023.03.17.23287396

**Authors:** Jason P. Block, Keith A. Marsolo, Kshema Nagavedu, L. Charles Bailey, Tegan K. Boehmer, Julia Fearrington, Aaron M. Harris, Nedra Garrett, Alyson B. Goodman, Adi V. Gundlapalli, Rainu Kaushal, Abel Kho, Kathleen M. McTigue, Vinit P. Nair, Jon Puro, Elizabeth Shenkman, Mark G. Weiner, Neely Williams, Thomas W. Carton, PCORnet^®^ Network Partners

## Abstract

**Background:** Prior studies have documented differences in the age, racial, and ethnic characteristics among patients with SARS-CoV-2 infection. However, little is known about how these characteristics changed over time during the pandemic and whether racial, ethnic, and age disparities evident early in the pandemic were persistent over time. This study reports on trends in SARS-CoV-2 infections among U.S. adults from March 1, 2020 to January, 31 2022, using data from electronic health records.

**Methods and Findings:** We captured repeated cross-sectional information from 43 large healthcare systems in 52 U.S. States and territories, participating in PCORnet^®^, the National Patient-Centered Clinical Research Network. Using distributed queries executed at each participating institution, we acquired information for all patients ≥ 20 years of age who were tested for SARS-CoV-2 (both positive and negative results), including care setting, age, sex, race, and ethnicity by month as well as comorbidities (assessed with diagnostic codes).

During this time period, 1,325,563 patients had positive (13% inpatient) and 6,705,868 patients had negative (25% inpatient) viral tests for SARS-CoV-2. Disparities in testing positive were present across racial and ethnic groups, especially in the inpatient setting. Compared to White patients, Black or African American and other race patients had relative risks for testing positive of 1.5 or greater in the inpatient setting for 12 of the 23-month study period. Compared to non-Hispanic patients, Hispanic patients had relative risks for testing positive in the inpatient setting of 1.5 or greater for 16 of 23. Ethnic and racial differences were present in emergency department and ambulatory settings but were less common across time than in inpatient settings. Trends in infections by age group demonstrated higher test positivity for older patients in the inpatient setting only for most months, except for June and July of 2020, April to August 2021, and January 2022. Comorbidities were common, with much higher rates among those hospitalized; hypertension (38% of patients SARS-CoV-2 positive vs. 29% for those negative) and type 2 diabetes mellitus (22% vs. 13%) were the most common.

**Conclusion and Relevance:** Racial and ethnic disparities changed over time among persons infected with SARS-CoV-2. These trends highlight potential underlying mechanisms, such as poor access to care and differential vaccination rates, that may have contributed to greater disparities, especially early in the pandemic. Monitoring data on characteristics of patients testing positive in real time could allow public health officials and policymakers to tailor interventions to ensure that patients and communities most in need are receiving adequate testing, mitigation strategies, and treatment.

---

Multiple studies have documented the substantial differences in characteristics of patients with SARS-CoV-2 early in the pandemic, with individuals of minority race or ethnicity and older age bearing a much higher burden of severe COVID-19 illness (1–4). The US Centers for Disease Control and Prevention (CDC) has provided detailed information about cases and deaths by demographic characteristics, over time, gathered from data provided by state and local health departments and reference laboratories (5). These data have shown that rates of deaths per 100,000 spiked among non-Hispanic Black individuals early in the pandemic, followed by a decline compared to other racial and ethnic groups in the Summer and Fall 2020 and another increase starting in early 2021. Similarly, death rates among Hispanic individuals have remained higher than non-Hispanic individuals throughout the pandemic. However, to date, there has been limited information about patient characteristics and disparities among COVID-19 patients across care settings over time, compared to non-COVID-19 patients, such as those testing negative for SARS-CoV-2. These comparisons can help determine whether characteristics of patients and temporal trends were unique to patients with COVID-19 or were consistent with trends among patients with other illnesses. Further, this information can foster understanding of pandemic dynamics, both for ongoing public health response and to help determine how best to prepare for future public health crises.

Analyzing trends over time across care settings requires availability of longitudinal information on large populations throughout the entire course of the pandemic with detailed information about individuals, which is typically not available from public health authorities and public health departments. Research networks fill this void by using granular data from many healthcare systems, providing real-world information on diverse populations, followed for long periods.

Funded by the Patient-Centered Outcomes Research Network^®^ (PCORI^®^), PCORnet^®^,the National Patient-Centered Clinical Research Network, is a network-of-networks that facilitates research using electronic health record (EHR), administrative health plan claims, and other healthcare data (6,7). PCORnet currently has participation from more than 60 data contributing sites (i.e., individual health systems) embedded within nine Clinical Research Networks (8). Using data from 23 months of the pandemic (March 2020 through January 2022) in 43 institutions participating in PCORnet, we describe demographic trends over time of one of the largest US national sample of patients testing positive or negative for SARS-CoV-2.

## Methods

### PCORnet^®^ Common Data Model

PCORnet^®^ operates as a distributed research network, where data are held locally behind institutional firewalls, with queries sent to institutions from a central PCORnet^®^ Coordinating Center. The participating sites utilize the PCORnet^®^ Common Data Model, in which each data contributing site maps its clinical and/or administrative claims data into a standardized representation (9). These data are normally updated quarterly by sites, coinciding with data quality curation cycles in which data are examined for conformance with the Common Data Model specifications as well as data plausibility and data completeness.

Prompted by the COVID-19 pandemic, PCORnet^®^ sites agreed to rapidly implement supplemental data extraction/curation procedures to allow for rapid capture of information on patients with COVID-19. Starting in early April 2020, sites began updating COVID-related data at least monthly (as often as daily) instead of quarterly. Rather than doing this for all patients (which requires considerable processing time and effort), PCORnet^®^ sites included a subset of all patients, including only those who had at least one International Classification of Diseases, 10^th^ revision, Clinical Modification (ICD-10-CM) diagnostic code for a potential viral illness, including COVID-19, or had a lab result for a SARS-CoV-2 test recorded beginning January 1, 2020 (**Supplemental Figure 1**) (10). Through January 31, 2022, this database included 16,317,253 individual patients 20+ years of age.

### Queries and Patient Characterization

From this larger population of patients, we used existing modular programs to characterize all patients tested for SARS-CoV-2 (**Supplemental Methods Note**). These programs return only aggregate data from sites. Submission, and use of these data are covered by a PCORnet-wide Data Sharing Agreement and individual site Institutional Review Board approvals for their PCORnet Common Data Model data. Using designated Logical Observation Identifiers Names and Codes (LOINC) (11), we captured information on patients with results of “positive,” “presumptive positive,” or “detected” on nuclear acid amplification (NAAT)/polymerase chain reaction (PCR) or antigen viral tests for SARS-CoV-2 (referred to as “testing positive” or “positive viral test” hereafter) as well as those with results of “negative (“testing negative” or “negative viral test”). Patients included in the negative viral test strata had only records of negative tests and no positive tests. We stratified patients based on the setting of their viral test: 1) all, 2) ambulatory, 3) emergency department, and 4) inpatient. The latter three categories were mutually exclusive. We assigned a care setting by determining the setting for encounters before and after the test was performed (1 day before and 16 days following). To make care settings exclusive, we removed patients from a lower care setting stratum if they had encounters in a higher care setting (for ambulatory strata, we assessed emergency and inpatient encounters) before and after their positive test (21 days before and 16 days following the date of the test) (**Supplemental Note**). Patients identified as receiving care in the emergency department included patients who were discharged directly from this setting and not admitted to the hospital within 16 days; those admitted directly from the emergency department or soon thereafter were characterized with the inpatient care setting cohort. One-third of patients did not have a care setting associated with their positive test, perhaps because many patients were tested in drive-thru testing settings that might not have registered an encounter; some of these tests might also have been ordered during telehealth or virtual visits that were not specifically assessed in our queries. We captured all information with respect to the date for an index viral laboratory test, which was the first positive test for patients testing positive and the first negative test for patients testing negative. All codes for case definitions, comorbidities, treatments, and labs were posted on GitHub.

### Demographics, Comorbidities, and Disease Severity

Using information available at the time of the test, we assessed demographic characteristics of patients (i.e., age group, sex, race, and ethnicity) using value sets defined in the PCORnet Common Data Model. Sex categories allowed for Male, Female, and Other. Several race categories (i.e., Multiple race, Other race, Native Hawaiian/Other Pacific Islander, and Alaskan Native/American Indian) were less common and typically <1% for all strata; these were grouped together under “Other Race.” Ethnicity was defined as Hispanic or not Hispanic, with a separate missing category. We assessed monthly trends by age group, race, and ethnicity by care setting. We assessed disparities by calculating monthly patient positivity as the total number of patients testing positive separately for each racial, ethnic and age group, divided by the total number of patients tested in that month by care setting. We calculated relative risks (RRs) by month as the percentage of patients testing positive by racial, ethnic, and age group divided by the percentage in a reference group (White; non-Hispanic; age 20-39 years, respectively for each assessment of patient characteristics); these calculations were done separately for race, ethnicity, and age. We noted that disparities were present when RRs were ≥1.5.

We included a detailed assessment of comorbidities, which were selected based on information presented in publications regarding the association of comorbidities with COVID-19 outcomes (12–15). Patients were considered to have a comorbidity if they had at least two ICD-9-CM or ICD-10-CM codes for the disease/condition recorded any time over a three-year period prior to the index date of their viral test. We also captured use of critical care procedure/billing Current Procedural Terminology (CPT) codes (99291, 99292) during this period and mechanical ventilation usage through presence of ICD-10-CM, Healthcare Common Procedure Coding System (HCPCS) and CPT codes any time from the index date of the SARS-CoV-2 test through 21 days after the index date. Lastly, we categorized weight status using the most recent height and weight available within the year prior to the index date. This study was reviewed by the Institutional Review Board of Harvard Pilgrim Health Care, Inc., which deemed it exempt from review because it was a public health surveillance project, conducted under the direction of a public health authority. This activity was reviewed by the CDC and was conducted consistent with applicable federal law and CDC policy.

## Results

### Characteristics of patients testing positive for SARS-CoV-2

The query captured aggregate data on 1,325,563 patients who tested positive for SARS-CoV-2 and 6,705,868 patients who tested negative from March 1, 2020 through January 31, 2022. Across the 43 sites, percent contributions to the overall sample ranged from <1% to 15% with 15 sites each contributing ≥ 3% of the total; these 15 sites included 73% of all patients testing positive for SARS-CoV-2. Of patients testing positive, 38% were treated in the ambulatory setting, 15% were seen and discharged from the emergency department, and 13% were hospitalized (**Table 1**). Among hospitalized patients, 10% and 19% were documented as receiving mechanical ventilation and critical care treatment, respectively. Test positivity among patients (not tests) was 17% overall and 15%, 20%, and 9% in the ambulatory, emergency department, and inpatient setting, respectively.

**Table 1:** Characteristics of patients positive for SARS-CoV-2 on polymerase chain reaction or antigen laboratory tests, March 1, 2020 – January 30, 2022, 43 PCORnet Data Contributing Institutions

|  | All SARS-CoV-2 + |  |  | SARS-CoV-2 +, Ambulatory |  |  | SARS-CoV-2 +, Emergency Department |  |  | SARS-CoV-2 +, Inpatient |  |  |
| --- | --- | --- | --- | --- | --- | --- | --- | --- | --- | --- | --- | --- |
|  | 1,325,563 |  |  | 499,370 |  |  | 202,241 |  |  | 172,673 |  |  |
|  | N | % | RR | N | % | RR | N | % | RR | N | % | RR |
| **Age (years)** |  |  |  |  |  |  |  |  |  |  |  |  |
| 20-<40 | 540,033 | 41% | Ref | 221,535 | 44% | Ref | 85,360 | 42% | Ref | 31,125 | 18% | Ref |
| 40-<55 | 346,074 | 26% | 1.0 | 138,259 | 28% | 0.9 | 53,762 | 27% | 1.0 | 33,280 | 19% | 1.7 |
| 55-<65 | 204,153 | 15% | 0.8 | 74,873 | 15% | 0.6 | 29,682 | 15% | 0.9 | 34,186 | 20% | 1.8 |
| 65-<75 | 135,503 | 10% | 0.6 | 43,829 | 9% | 0.5 | 18,926 | 9% | 0.7 | 34,580 | 20% | 1.7 |
| 75-<85 | 70,592 | 5% | 0.6 | 17,017 | 3% | 0.4 | 10,162 | 5% | 0.6 | 25,748 | 15% | 1.9 |
| 85+ | 29,208 | 2% | 0.7 | 3,857 | 1% | 0.4 | 4,349 | 2% | 0.5 | 13,754 | 8% | 2.2 |

| Sex |  |  |  |  |  |  |  |  |  |  |  |  |
| --- | --- | --- | --- | --- | --- | --- | --- | --- | --- | --- | --- | --- |
| *Female* | 736,587 | 56% | Ref | 284,413 | 57% | Ref | 112,905 | 56% | Ref | 86,107 | 50% | Ref |
| *Male* | 588,459 | 44% | 1.1 | 214,851 | 43% | 1.0 | 89,312 | 44% | 1.0 | 86,545 | 50% | 1.4 |
| *Other<sup>6</sup>* | 511 | 0% | * | 102 | 0% | * | 24 | 0% | * | 20 | 0% | * |
| Race |  |  |  |  |  |  |  |  |  |  |  |  |
| *Asian* | 37,423 | 3% | 0.9 | 13,153 | 3% | 0.9 | 4,935 | 2% | 1.0 | 5,594 | 3% | 1.2 |
| *Black or African American* | 214,540 | 16% | 1.1 | 62,218 | 12% | 1.0 | 56,796 | 28% | 1.2 | 39,384 | 23% | 1.5 |
| *White* | 831,180 | 63% | Ref | 343,881 | 69% | Ref | 100,859 | 50% | Ref | 91,793 | 53% | Ref |
| *Other* | 126,993 | 10% | 1.1 | 38,029 | 8% | 1.1 | 32,338 | 16% | 1.3 | 26,716 | 15% | 1.8 |
| *Missing* | 115,427 | 9% | 1.0 | 42,089 | 8% | 1.2 | 7,313 | 4% | 1.1 | 9,186 | 5% | 1.5 |

| Hispanic Ethnicity |  |  |  |  |  |  |  |  |  |  |  |  |
| --- | --- | --- | --- | --- | --- | --- | --- | --- | --- | --- | --- | --- |
| *Yes* | 217,931 | 16% | 1.5 | 69,250 | 14% | 1.5 | 45,620 | 23% | 1.5 | 33,254 | 19% | 1.8 |
| *No* | 937,127 | 71% | Ref | 370,700 | 74% | Ref | 139,387 | 69% | Ref | 126,686 | 73% | Ref |
| *Other/Missing* | 170,505 | 13% | 0.9 | 59,420 | 12% | 0.9 | 17,234 | 9% | 1.2 | 12,733 | 7% | 1.2 |
| Month |  |  |  |  |  |  |  |  |  |  |  |  |
| *March 2020* | 19,823 | 1% |  | 4,121 | 1% |  | 4,007 | 2% |  | 7,157 | 4% |  |
| *April 2020* | 39,561 | 3% |  | 10,117 | 2% |  | 6,257 | 3% |  | 13,462 | 8% |  |
| *May 2020* | 25,207 | 2% |  | 9,416 | 2% |  | 4,989 | 2% |  | 4,849 | 3% |  |
| *June 2020* | 36,650 | 3% |  | 16,635 | 3% |  | 6,045 | 3% |  | 4,559 | 3% |  |
| *July 2020* | 60,330 | 5% |  | 28,146 | 6% |  | 8,698 | 4% |  | 7,299 | 4% |  |
| *August 2020* | 37,832 | 3% |  | 18,200 | 4% |  | 4,552 | 2% |  | 4,915 | 3% |  |
| *September 2020* | 34,122 | 3% |  | 19,109 | 4% |  | 3,247 | 2% |  | 3,897 | 2% |  |
| *October 2020* | 60,474 | 5% |  | 26,828 | 5% |  | 6,386 | 3% |  | 6,178 | 4% |  |
| *November 2020* | 125,513 | 9% |  | 48,605 | 10% |  | 13,789 | 7% |  | 11,149 | 6% |  |
| *December 2020* | 131,017 | 10% |  | 46,042 | 9% |  | 14,255 | 7% |  | 14,713 | 9% |  |
| *January 2021* | 97,096 | 7% |  | 31,522 | 6% |  | 12,466 | 6% |  | 14,481 | 8% |  |
| *February 2021* | 39,816 | 3% |  | 12,239 | 2% |  | 5,749 | 3% |  | 7,972 | 5% |  |
| *March 2021* | 32,802 | 2% |  | 9,016 | 2% |  | 5,905 | 3% |  | 6,889 | 4% |  |
| *April 2021* | 32,097 | 2% |  | 8,490 | 2% |  | 6,735 | 3% |  | 6,499 | 4% |  |
| *May 2021* | 15,190 | 1% |  | 3,993 | 1% |  | 3,631 | 2% |  | 3,488 | 2% |  |
| *June 2021* | 8,414 | 1% |  | 2,555 | 1% |  | 1,815 | 1% |  | 2,118 | 1% |  |
| *July 2021* | 22,919 | 2% |  | 7,511 | 2% |  | 6,776 | 3% |  | 4,420 | 3% |  |
| *August 2021* | 62,668 | 5% |  | 22,775 | 5% |  | 14,955 | 7% |  | 9,832 | 6% |  |
| *September 2021* | 52,779 | 4% |  | 20,113 | 4% |  | 10,079 | 5% |  | 7,964 | 5% |  |
| *October 2021* | 33,004 | 2% |  | 12,400 | 2% |  | 5,478 | 3% |  | 5,130 | 3% |  |
| *November 2021* | 35,651 | 3% |  | 13,750 | 3% |  | 6,220 | 3% |  | 4,654 | 3% |  |
| *December 2021* | 109,629 | 8% |  | 42,197 | 8% |  | 21,346 | 11% |  | 7,857 | 5% |  |
| *January 2022* | 212,969 | 16% |  | 85,590 | 17% |  | 28,861 | 14% |  | 13,191 | 8% |  |

Black or African American patients were 16% of all patients testing positive but were 12%, 28%, and 23% of patients testing positive in the ambulatory, emergency department, and inpatient settings, respectively (**Table 1**). Other Race patients were 10% of all patients testing positive but were 8%, 16%, and 15% in the ambulatory, emergency department, and inpatient settings. The RR for testing positive for Black or African American patients vs. White patients was 1.0, 1.2, and 1.5 across the care settings, and RR for Other Race patients was 1.1, 1.3, and 1.8 (**Table 1**). Compared to non-Hispanic patients, the RR for testing positive in the ambulatory, emergency department, and inpatient settings for Hispanic patients was 1.5, 1.5, and 1.8 (**Table 1**).

Overall, more patients testing positive were in the 20 to 39 years age group (41%) than other age groups, primarily driven by patients testing positive in the ambulatory setting (**Table 1**). Patients 20-39 years of age comprised the largest proportion of patients testing positive in the ambulatory and emergency department settings, but 55-74 year old patients comprised the largest proportion in the inpatient setting (Table 1). Compared to patients who were 20-39 years, the RR for testing positive in the inpatient setting was 1.7 for patients 40 to <55 years, 1.8 for patients 55 to <65 years, 1.7 for patients 65 to <75 years, 1.9 for patients 75-<85 years, and 2.2 for patients 85+ years. Women comprised a larger proportion of patients testing positive overall (56%), with men more likely to test positive only in the inpatient setting (RR 1.4).

### Trends by race, ethnicity, and age

Disparities by race were most apparent during the early phase of the pandemic, especially in the inpatient setting. Among patients hospitalized with SARS-CoV-2 in March and April 2020, Black or African American, Other race, and White patients comprised nearly an equal proportion of patients (**Supplemental Figure 2**). Black or African American patients still comprised more than 20% of patients hospitalized for 17 of the 23 months examined. When we calculated relative risks by month, we demonstrated that hospitalized Black or African American patients were twice as likely to test positive for SARS-CoV-2 compared to White hospitalized patients from April through August 2020 and 1.5 times as likely to tested positive for 12 of the 23 months, with the lowest RRs from September 2020 to January 2021 and again from June to November 2021 (**Figure 1**). Patients with “Other Race” had similar patterns when compared to patients of White race, with relative risks of 1.5 or greater for 12 of the 23 months. Patients with Missing or Asian race followed a similar pattern to Black and Other Race groups early in the pandemic and from December 2020 through April 2021. These disparities were similar in the emergency department, with lower relative risks than the inpatient setting (**Supplemental Figure 3**). Disparities also were noted in the ambulatory setting but only in the early months of the pandemic (**Figure 1**).

**Figure 1:**
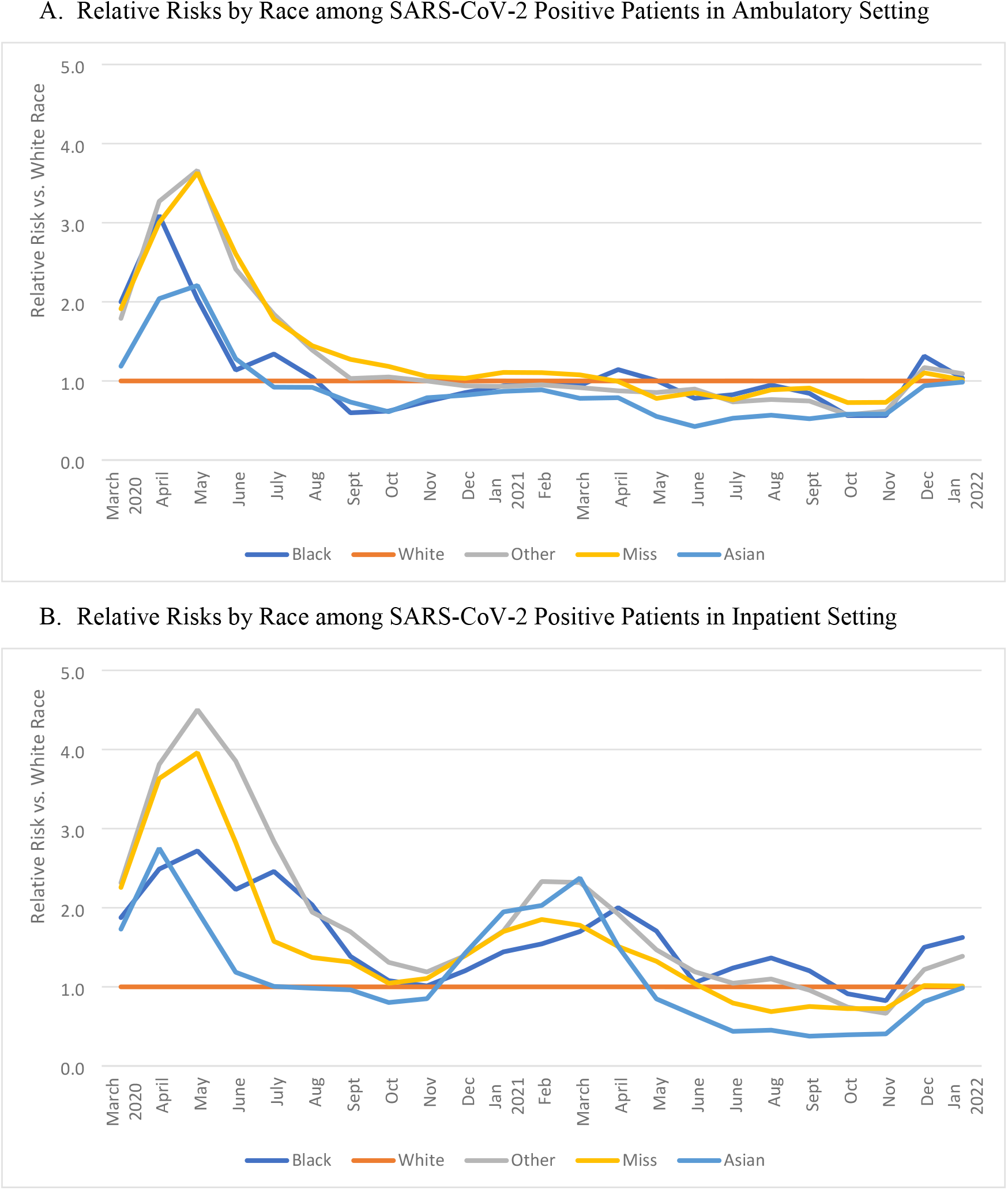
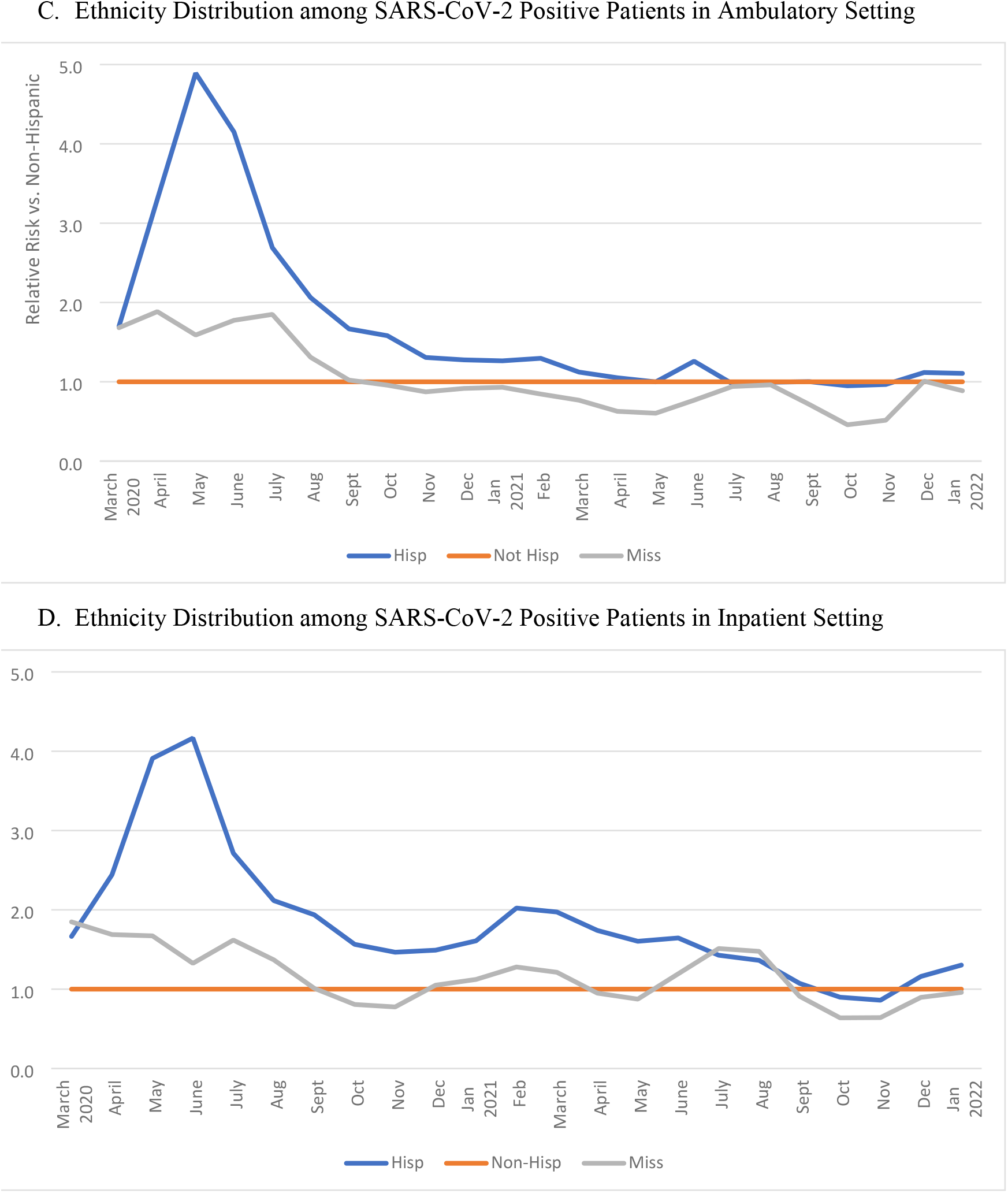
Monthly relative risks for testing positive for SARS-CoV-2 by race and ethnicity and care setting, March 1, 2020 to January 31, 2022. These figures show the relative risk for testing positive by month for Black or African American, Asian, Other Race or Missing Race compared to White race as a reference in the (A) ambulatory or (B) inpatient setting and those who were Hispanic, or had Missing Hispanic ethnicity compared to Not Hispanic as a reference in the (C) ambulatory or (D) inpatient setting. Care settings are mutually exclusive. Proportions of patients testing positive by care setting are available in Supplemental Figures 2 and 4; relative risks for testing positive in the emergency department are available in Supplemental Figures 3 and 5. Abbreviations: Hisp-Hispanic; Not Hisp-Not Hispanic; Miss-Missing.

Trends were similar when examined by ethnicity, with a disproportionate number of patients with Hispanic ethnicity testing positive across all care settings; these disparities persisted through mid-2021 in the emergency department in inpatient settings (**Figure 1; Supplemental Figures 4-5**). Compared to non-Hispanic patients, Hispanic patients had relative risks for testing positive in the inpatient setting of 1.5 or greater for 16 months.

Trends in infections by age demonstrated consistently higher test positivity for 20-39-year-olds in the ambulatory setting (**Figure 2, Supplemental Figures 6-7**). In the emergency department, younger patients were more likely to test positive than older patients in most months except for March to April 2020 (**Supplemental Figure 7**). Patients in older age groups (55 years and higher) were more likely to test positive in the inpatient setting than patients who were 20-39 years, with RRs ≥1.5 consistently except in June-July 2020, April-July 2021, and December 2021-January 2022 (**Figure 2, Supplemental Figure 6**); patients who were 40 to 54 years had RRs ≥1.5 for nearly all months of the pandemic compared to patients 20-39 years, except December 2021-January 2022.

**Figure 2:**
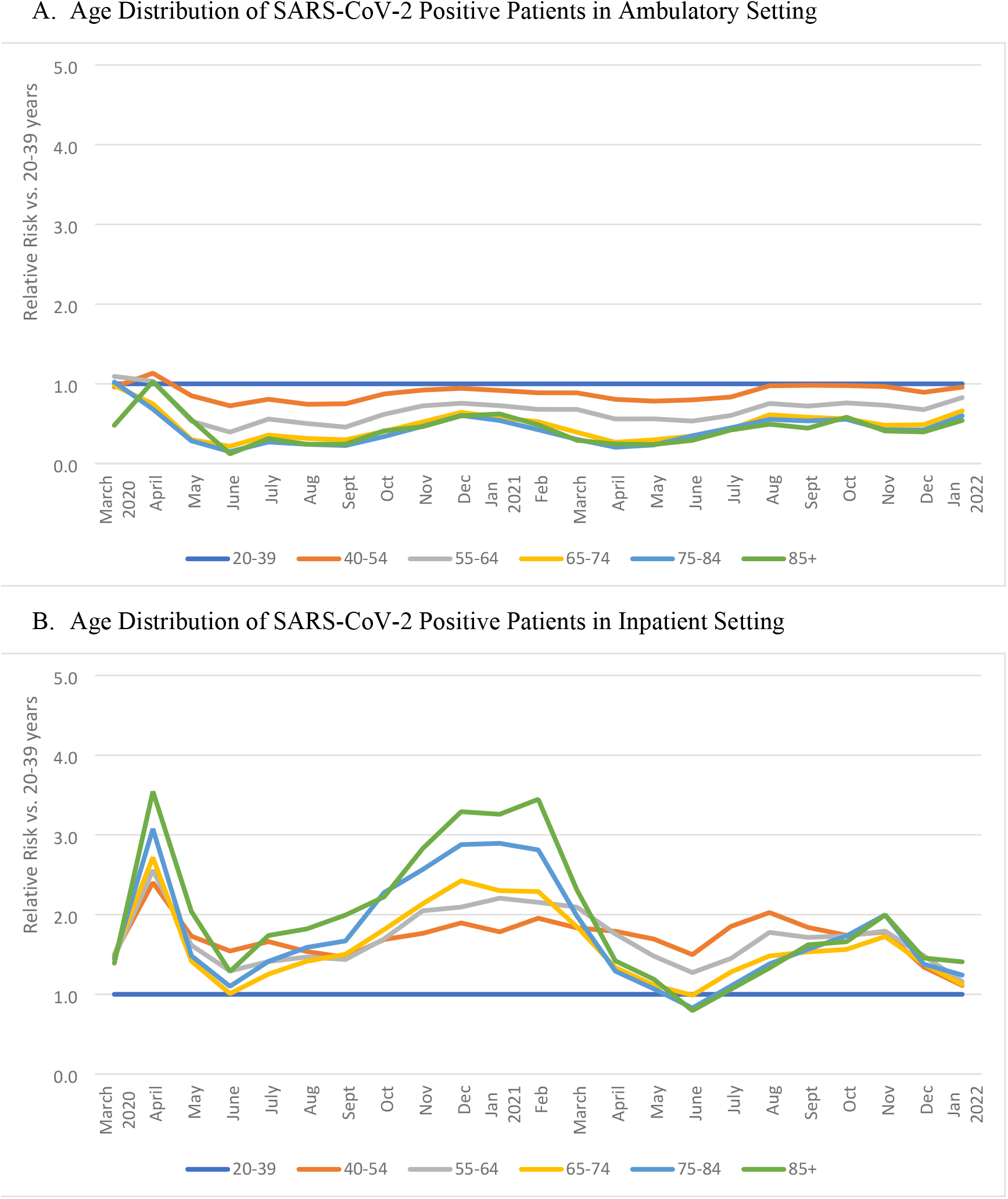
Monthly relative risks for testing positive for SARS-CoV-2 by age group and care setting, March 1, 2020 to January 31, 2022. These figures show the relative risks for testing positive by month and age group in the (A) ambulatory and (B) inpatient setting; reference was 20-39 years of age. Proportions of patients testing positive by care setting are available in Supplemental Figure 6; relative risks for testing positive in the emergency department are available in Supplemental Figure 7. Care settings are mutually exclusive.

### Comorbidities

Among patients testing positive for SARS-CoV-2, hypertension was the most common comorbidity (19% for all patients) followed by type 2 diabetes (DM2) (9%), chronic pulmonary disease (7%), and mental health disorders (8%); 7% of patients had hypertension and either Type 1 (T1DM) or T2DM (**Figure 3; Supplemental Table 3**). These diagnoses were more common among patients with SARS-CoV-2 treated in the inpatient setting, with hypertension present in 38% and DM2 in 22% (19% had hypertension and either DM1 or DM2) of inpatients. The prevalence of obesity also was high at 46% among all patients testing positive but was similar across care settings (45% to 48%). Patients testing negative while inpatient had a lower prevalence of hypertension (29% vs. 38%), diabetes (13% vs. 22%), and obesity (41% vs. 47%) compared with those testing positive (**Supplemental Table 4**); prevalence was similar for coronary artery disease (10% vs. 13%), mental health disorders (11% vs. 12%), or chronic pulmonary disease (11% vs. 14%).

**Figure 3:**
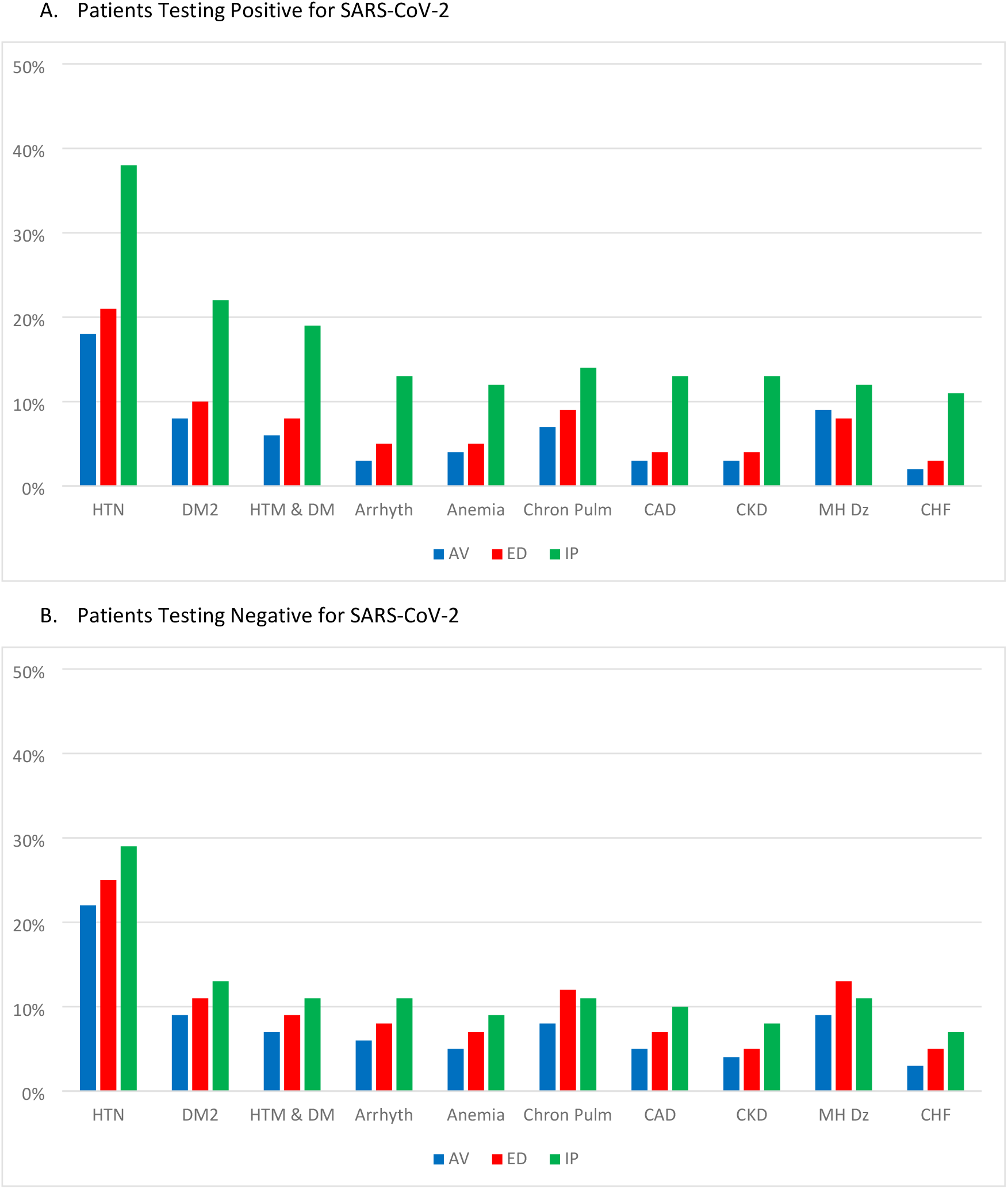
Comorbidities for patients with positive viral tests for SARS-CoV-2 across care settings, March 1, 2020 to January 31, 2022. These figures show prevalence of various comorbidities for patients with positive viral tests for SARS-CoV-2 across care settings. Each condition includes a broad capture of ICD-9-CM and ICD-10-CM codes, with codes lists available on Github. For example, mental health codes include those for a large number associated with psychotic and mood disorders. Care settings were mutually exclusive. Abbreviations: AV-ambulatory visit; ED-emergency department; IP-inpatient or hospitalized; HTN-hypertension; DM2-diabetes mellitus type 2, DM-diabetes mellitus Types 1 and2, Arrhyth-Arrhythmia; Chron Pulm-Chronic Pulmonary Disease; CAD-coronary artery disease; CKD-chronic kidney disease; MH Dis-mental health disease; CHF-congestive heart failure.

## Discussion

Using real world electronic health care data, we examined a large population of patients with SARS-CoV-2 infection, using information on 1.3 million in 43 institutions testing positive for SARS-CoV-2 infection and 6.7 million who tested negative without any positive tests through January 2022. Unlike most other studies, we compared patients across care settings, with rich demographic and comorbidity data by ambulatory, emergency department, and inpatient settings; we also compared patients who tested positive to those who tested negative. Trends by race, ethnicity and age revealed important information about health inequities during the COVID-19 pandemic. While variable over time, individuals who were Black or African American or Hispanic were more likely to test positive compared to White and non-Hispanic patients in the inpatient setting for most months.

Beyond basic demographic trends in cases and death overall, as reported by public health authorities, few studies have reported demographic trends in infections. Studies that have reported on these trends have covered short-term trends early in the pandemic; this study covers 23 months of the pandemic. Using data from a national reference laboratory (ARUP Laboratories, with nearly half of the sample of 19,320 patients from Utah), investigators at the US Centers for Disease Control and Prevention (CDC) found a changing age distribution of patients infected with SARS-CoV-2, with a decline in median age in June and July 2020 of 35.8 years compared to 40.5 in March and April 2020 (16). From May to August 2020, state electronic laboratory reporting data from 37 states showed similar trends by age, with 20-to-29-year-olds having the highest incidence of SARS-CoV-2 among all age groups from June through August (4). This increase in incidence among young people continued through early September, as 18- to 22-year-olds started to return to colleges (3). In our study, we found peaks in test positivity of young people during the Summer of 2020 and 2021.

In our study, racial and ethnic disparities in infections, especially among those hospitalized, were evident throughout the pandemic but were most evident early in the pandemic. Disparities by race, defined by test positivity, in the inpatient setting were highest early in the pandemic and during the Winters of 2020/21 and 2021/22. Ethnic disparities were persistent for most of the pandemic in the inpatient setting, with a higher proportion of Hispanic individuals testing positive when hospitalized compared to non-Hispanic patients; this trend ended in Fall 2021.

Other studies have found similar patterns for shorter periods. In COVID-19 mortality data from the National Vitals Statistics System, disparities were evident from May to August 2020, with some differences over time (1). Overall, 19% of individuals who died from COVID-19 were Black or African American, with a reduction in proportion from 20% in May 2020 to 17% in August; 24% of all people who died were Hispanic, with an increase from 16% to 26% during this same time period. From January 26 to October 3, 2020, there were nearly 300,000 excess deaths in the United States; this rate summarizes all-cause mortality compared to expected rates from prior years, in this case 2015 to 2019 (2). Substantial disparities were present. The excess death rate was 11.9% for Whites but 29% to 37% for racial minorities and 53% for Hispanic persons; this was highest in March and April 2020 with a second peak among Hispanics in July and August 2020.

Among all conditions assessed, we found higher prevalence of hypertension, type 2 diabetes mellitus, and obesity among adults treated in the inpatient setting with SARS-CoV-2 compared to those testing negative. These differences could reflect the demographic differences, including age, found between those testing negative and positive or an increased risk of severe disease among patients with these conditions. Diabetes has been found to be an independent risk factor for severe COVID-19, including mortality, likely secondary to underlying physiologic mechanisms, such as immune effects from diabetes and a higher affinity for cellular binding of the virus in patients with diabetes (17–20). The high prevalence of diabetes especially in our study is further evidence of a greater burden of severe SARS-CoV-2 in this population.

COVID-19 surveillance in PCORnet provides several unique opportunities. Obtaining real-world, comprehensive information on a diverse, national population of patients with COVID-19 is critical to help guide policymakers as well as clinical and public health leaders regarding planning for future COVID-19 response. The network infrastructure and large samples allows for creation of research-ready cohorts and in-depth assessment of patients, based on demographic or clinical characteristics. Analyses can be done in a distributed fashion without sharing patient-level data, including predictive modeling, and aggregation across diverse sites can increase generalizability. Large sample sizes with granular clinical information will be increasingly important to capture information on patient subgroups, defined by demographics or less common presentations or outcomes of COVID-19, such as rare complications of SARS-CoV-2. Prior PCORnet surveillance studies have examined disparities in use of monoclonal antibodies (21), the occurrence of post-acute sequelae of COVID-19 (22), and the incidence of myocarditis and pericarditis after SARS-CoV-2 infection and COVID-19 vaccination (23). The investment in changing the processes for data management – moving to frequent updates for patients with respiratory illnesses from a quarterly update – prepared the network to make ongoing contributions to the investigation of COVID-19. Other national initiatives, such as The National COVID Cohort Collaborative (N3C) are gathering similar information from institutions, with patient-level data extracts, to allow for varied research on COVID-19 over time (24).

### Limitations

Several limitations are inherent to working with structured healthcare data. First, electronic health records and administrative claims data may not have complete data on some patients, especially patients tested and treated entirely outside of a PCORnet health system or outside of the health plan insurance coverage. In such cases, the PCORnet Common Data Model may not have any information about a SARS-CoV-2 infection or inadequate information on chronic diseases and medication use. For example, some institutions had less data on symptomatic ambulatory patients, who may have been referred to outside facilities for testing; other institutions did their own testing for symptomatic ambulatory patients. Most importantly, missing information could have led to misclassification, such as when a patient only had negative tests in the healthcare system but tested positive at an outside institution or on a home test, results that would not be captured in the PCORnet Common Data Model. If missingness was differential by demographic characteristics, proportions reported could be inaccurate.

Second, even for patients who received all care within a PCORnet institution, missing data were still possible, as we found for variables such as race, weight status, and care setting. If missingness was non-random, which could be the case if data were less available on patients of certain race categories, our results could provide some incomplete conclusions. Third, the results we report are descriptive and unadjusted for potential confounders. We discovered higher rates of several chronic conditions among patients with SARS-CoV-2 compared to those without; these differences might be explained by demographic or age differences. We required at least two ICD-10-CM codes for chronic conditions, which may have led to an undercount, especially for those patients who were not seen often in the healthcare system prior to their SARS-CoV-2 test. Fourth, we report differences based on race/ethnicity information available in the EHR. We do not have further information in the Common Data Model, such as socioeconomic status or other information that underlie disparities, such as exposure to racism or other social determinants. We also do not have access to payer status, which could provide some information on disparities by type of insurance, such as governmental or private. Finally, PCORnet does not allow for representative estimates of overall COVID-19 cases across the U.S. Because PCORnet has a fixed number of institutions and is not designed to be representative of the U.S. population, the prevalence of cases across states in PCORnet is not equivalent to the actual state prevalence of patients who have been tested for SARS-CoV-2.

## Conclusion

PCORnet has captured information on one of the largest samples to date of patients tested for SARS-CoV-2 infection. Racial and ethnic minority patients have borne a disproportionate burden of the pandemic, making up a larger proportion of those infected than other groups, especially for those hospitalized. Monitoring these data in real time could allow public health officials and policymakers to develop messaging and targeted programs for these communities to ensure that they are receiving adequate testing, mitigation strategies, and treatment. Patients diagnosed with COVID-19 were found to have a high burden of underlying conditions, especially among those hospitalized.

## Data Availability

This data was captured using distributed queries of healthcare system clinical data. We only have aggregate data and not the source data available for this publication. All relevant data for this publication is included in the manuscript itself. Requests for further data relevant to this publication are available through the PCORnet Front Door: https://pcornet.org/front-door/.

## Abbreviations

EHR: electronic health record
CDM: Common Data Model
ETL: extract-transfer-load procedure to move data from a clinical data warehouse to the PCORnet Common Data Model
Arrhyth: arrhythmia
CAD: coronary artery disease
CHF: congestive heart failure
CKD: chronic kidney disease
Chron pulm: chronic pulmonary disease
Coag: Coagulopathy
DM2: type 2 diabetes mellitus
HTN: hypertension
Mental: mental health disorder
Amb: ambulatory care setting
ED: emergency department
Inpt: inpatient
Vent: inpatient with procedure codes indicating use of mechanical ventilation
conv plasma: convalescent plasma
dex: dexamethasone
HCQ: hydroxychloroquine
remdes: remdesivir
steroids: corticosteroids other than dexamethasone
tocili: tocilizumab

## Acknowledgement

This study was funded through a cooperative agreement to the Task Force for Global Health, through the US Centers for Disease Control and Prevention: COVID-19 Electronic Healthcare Data Initiative. Each of the institutions received funding through this agreement.

The Clinical Research Networks and Coordinating Center reported in this publication are Network Partners in PCORnet, the National Patient-Centered Clinical Research Network. PCORnet has been developed with funding from the Patient-Centered Outcomes Research Institute (PCORI). The Network Partners’ participation in PCORnet is funded through the following PCORI Awards: Coordinating Center (PCORI-CC2-Duke-2016); ADVANCE (RI-CRN-2020-001); CaPriCORN (RI-CRN-2020-002); GPC (RI-CRN-2020-003); HealthCore (HP-1510-32545); Insight (RI-CRN-2020-004); OneFlorida (RI-CRN-2020-005); PaTH (RI-CRN-2020-006); PEDSnet (RI-CRN-2020-007); REACHnet (RI-CRN-2020-008); and, STAR (RI-CRN-2020-009). Authors report no competing interests associated with this publication.

The funder provided support in the form of salaries or stipends for all authors. As the funding organization of the network infrastructure for PCORnet, PCORI participates in discussions and high-level decision-making about the network and its strategic priorities. With this participation, PCORI provided support and approval for the initiative to alter the PCORnet infrastructure and support queries related to COVID-19. However, PCORI but had no specific role in the study design, data collection and analysis, decision to publish, or preparation of the manuscript. The specific roles of these authors are articulated in the ‘author contributions’ section.

The findings and conclusions in this report are those of the authors and do not necessarily represent the official position of CDC or the US Public Health Service.

## Collaborative Authors: *PCORnet*^®^ *Network Partners*

Faraz S. Ahmad, MD, MS, Northwestern University Feinberg School of Medicine; Saul Blecker, NYU Grossman School of Medicine; H. Ryan Carnahan, PharmD, MS, The University of Iowa College of Public Health, Department of Epidemiology; Olveen Carrasquillo, MD, MPH, University of Miami Miller School of Medicine; Bernard P. Chang, MD, PhD, Columbia University Irvine Medical Center; Katherine Chung-Bridges, MD, MPH, Health Choice Network; Lindsay G. Cowell, MS, PHD, UT Southwestern Medical Center; Janis L. Curtis, MSPH, MA, Duke University School of Medicine; Christine Draper, Department of Population Medicine, Harvard Pilgrim Health Care Institute, Harvard Medical School; Soledad A. Fernandez, PhD, The Ohio State University; Christopher B. Forrest, MD, PhD, Children’s Hospital of Philadelphia; Nidhi Ghildayal, PhD, Department of Population Medicine, Harvard Pilgrim Health Care Institute, Harvard Medical School; David A Hanauer, MD, MS, University of Michigan; Rachel Hess, MD, MS, University of Utah; Benjamin D. Horne, PhD, MStat, MPH, Intermountain Medical Center Heart Institute, Stanford University School of Medicine; Philip Giordano, MD, FACEP, Orlando Health, Inc.; William Hogan, MD, MS, University of Florida; Wenke Hwang, PhD, Penn State University College of Medicine; Harold P. Lehmann MD, PhD, Johns Hopkins School of Medicine; Abel Kho, MD, MS, Northwestern University; Scott Mackey, DO, LCMC Health; Kenneth H. Mayer MD, Fenway Health; Narayana Mazumder, MS, Allina Health; James C. McClay, MD, University of Nebraska Medical Center; J. Greg Merritt, PhD, Patient is Partner, LLC; Abu Saleh Mohammad Mosa, Phd, MS, FAMIA, University of Missouri; Samyuktha Nandhakumar, MS, North Carolina Translational and Clinical Sciences Institute, UNC School of Medicine; Bridget Nolan, Department of Population Medicine, Harvard Pilgrim Health Care Institute; Jihad S. Obeid, MD, FAMIA, Medical University of South Carolina; Brian Ostasiewski, Wake Forest School of Medicine; Anuradha Paranjape, MD, MPH, Lewis Katz School of Medicine at Temple University; Lav Parshottambhai Patel, BS, MS, University of Kansas Medical Center; Patricia S. Robinson, PhD, APRN, AdventHealth Research Institute; Alexander Stoddard, MS, CTSI, Medical College of Wisconsin; William E. Trick, Cook County Health; Russell L. Rothman, MD, MPP, Vanderbilt University Medical Center; Neely Williams, MDiv, EdD

## Captions for Supporting Information

### S Methods Note

Describes process of code list generation, COVID-19 code-based case definitions, and sensitivity analyses for the assessment of comorbidities

**S1 Table: Responding PCORnet Healthcare Institutions**

**S2 Table: Characteristics of Patients Testing Negative for SARS-CoV-2 with No Positive Tests, March 1, 2020 to January 31, 2022**

**S3 Table: Comorbidities among Patients SARS-CoV-2 +, by Care Setting**

**S4 Table: Comorbidities among Patients SARS-CoV-2 -, by Care Setting**

### S Figures

**S Fig 1. PCORnet Common Data Models, including standard and COVID-19 Common Data Model**.

The PCORnet Common Data Model includes information collected through various clinical data systems, including electronic health records (EHR) and health insurance claims or pharmacy dispensing records. Once data are entered by the user, systems transfer data to a data warehouse that is unique to their own health system and system vendor (e.g., Epic, Cerner). The typical process for populating the PCORnet Common Data Model is through an extraction-transfer-loading (ETL) procedure that allows for transformation of raw information into a form that is compliant with the Common Data Model specifications. This transformation, which is done by each individual institution in keeping with PCORnet’s federated data infrastructure, allows for consistency across health systems and centralized querying of data, and the PCORnet ETL and Common Data Model data refresh occurs quarterly. The PCORnet COVID-19 Common Data Model utilizes a similar process but rather than including information on all patients, it includes information only on patients who have had at least one respiratory illness diagnostic code or a lab test ordered for SARS-CoV-2 since January 1, 2020. This process occurs frequently (as often as daily) and includes data with lag times between 1 day to 7 days, depending on the site.

**S Fig 2. Monthly Proportion of Patients Testing Positive for SARS-CoV-2 by Race and Care Setting**

These figures show the proportion of patients by month who were White, Black or African American, Asian, Other Race or Missing Race in the (A) Ambulatory, (B) Emergency Department, (C) Inpatient, and (D) Non-Emergency or Inpatient Setting (includes all patients except for those who were directly connected to an emergency department or inpatient encounter). We included (D) to account for the approximately 1/3 of patients who had no care setting connected to their positive test; patients may not have had a care setting if they had a test done in a drive-thru setting without an encounter or if the test was ordered in a virtual care setting.

**S Fig 3. Monthly relative risks for testing positive for SARS-CoV-2 by Race in the Emergency Department, March 1, 2020 to January 31, 2022**.

These figures show the relative risks for testing positive by month and racial group in the emergency department; reference was White Race. Care settings are mutually exclusive.

**S Fig 4. Monthly Proportion of Patients Testing Positive for SARS-CoV-2 by Ethnicity and Care Setting**

These figures show the proportion of patients by month who were Hispanic, Not Hispanic, or had Missing Hispanic ethnicity information in the (A) Ambulatory, (B) Emergency Department, (C) Inpatient, and (D) Non-Emergency or Inpatient Setting (includes all patients except for those who were directly connected to an emergency department or inpatient encounter). We included (B) to account for the approximately 1/3 of patients who had no care setting connected to their positive test; patients may not have had a care setting if they had a test done in a drive-thru setting without an encounter or if the test was ordered in a virtual care setting.

**S Fig 5. Monthly relative risks for testing positive for SARS-CoV-2 by Ethnicity in the Emergency Department, March 1, 2020 to January 31, 2022**.

These figures show the relative risks for testing positive by month and ethnicity group in the emergency department; reference was White Race. Care settings are mutually exclusive.

**S Fig 6. Monthly Proportion of Patients Testing Positive for SARS-CoV-2 by Age and Care SettingSARS-CoV-2 +, Patients in Emergency Departments and Patients in the Non-Emergency or Inpatient Settings, Age by Month**

These figures show the proportion of patients by month and age group in the (A) Ambulatory, (B) Emergency Department, (C) Inpatient, and (D) Non-Emergency or Inpatient Setting (includes all patients except for those who were directly connected to an emergency department or inpatient encounter). We included (B) to account for the approximately 1/3 of patients who had no care setting connected to their positive test; patients may not have had a care setting if they had a test done in a drive-thru setting without an encounter or if the test was ordered in a virtual care setting.

**S Fig 7. Monthly relative risks for testing positive for SARS-CoV-2 by Age in the Emergency Department, March 1, 2020 to January 31, 2022**.

These figures show the relative risks for testing positive by month and age group in the emergency department; reference was 20-39-year-olds. Care settings are mutually exclusive.

## References

1. Gold JAW, Rossen LM, Ahmad FB, Sutton P, Li Z, Salvatore PP, et al. Race, Ethnicity, and Age Trends in Persons Who Died from COVID-19 - United States, May-August 2020. MMWR Morbidity and mortality weekly report. 2020;69(42):1517–21.

2. Rossen LM, Branum AM, Ahmad FB, Sutton P, Anderson RN. Excess Deaths Associated with COVID-19, by Age and Race and Ethnicity - United States, January 26-October 3, 2020. MMWR Morbidity and mortality weekly report. 2020;69(42):1522–7.

3. Salvatore PP, Sula E, Coyle JP, Caruso E, Smith AR, Levine RS, et al. Recent Increase in COVID-19 Cases Reported Among Adults Aged 18-22 Years - United States, May 31-September 5, 2020. MMWR Morbidity and mortality weekly report. 2020;69(39):1419–24.

4. Boehmer TK, DeVies J, Caruso E, van Santen KL, Tang S, Black CL, et al. Changing Age Distribution of the COVID-19 Pandemic - United States, May-August 2020. MMWR Morbidity and mortality weekly report. 2020;69(39):1404–9.

5. CDC COVID data tracker 2021 [Available from: https://covid.cdc.gov/covid-data-tracker/#demographicsovertime.

6. Collins FS, Hudson KL, Briggs JP, Lauer MS. PCORnet: turning a dream into reality. Journal of the American Medical Informatics Association : JAMIA. 2014;21(4):576–7.

7. Fleurence RL, Curtis LH, Califf RM, Platt R, Selby JV, Brown JS. Launching PCORnet, a national patient-centered clinical research network. Journal of the American Medical Informatics Association : JAMIA. 2014;21(4):578–82.

8. Forrest C, McTigue K, Hernandez A, Cohen L, Cruz H, Haynes K, et al. PCORnet 2020: current state, accomplishments, and future directions. Submitted for publication. 2020.

9. Greene DN, Jackson ML, Hillyard DR, Delgado JC, Schmidt RL. Decreasing median age of COVID-19 cases in the United States-Changing epidemiology or changing surveillance? PloS one. 2020;15(10):e0240783.

10. Qualls LG, Phillips TA, Hammill BG, Topping J, Louzao DM, Brown JS, et al. Evaluating Foundational Data Quality in the National Patient-Centered Clinical Research Network (PCORnet(R)). EGEMS (Washington, DC). 2018;6(1):3.

11. International Classification of Diseases, Tenth Revision, Clinical Modification (ICD-10-CM) 2020 [Available from: https://www.cdc.gov/nchs/icd/icd10cm.htm.

12. Fu L, Wang B, Yuan T, Chen X, Ao Y, Fitzpatrick T, et al. Clinical characteristics of coronavirus disease 2019 (COVID-19) in China: A systematic review and meta-analysis. The Journal of infection. 2020.

13. Guan WJ, Ni ZY, Hu Y, Liang WH, Ou CQ, He JX, et al. Clinical Characteristics of Coronavirus Disease 2019 in China. The New England journal of medicine. 2020;382(18):1708–20.

14. Liang WH, Guan WJ, Li CC, Li YM, Liang HR, Zhao Y, et al. Clinical characteristics and outcomes of hospitalised patients with COVID-19 treated in Hubei (epicenter) and outside Hubei (non-epicenter): A Nationwide Analysis of China. The European respiratory journal. 2020.

15. Richardson S, Hirsch JS, Narasimhan M, Crawford JM, McGinn T, Davidson KW, et al. Presenting Characteristics, Comorbidities, and Outcomes Among 5700 Patients Hospitalized With COVID-19 in the New York City Area. Jama. 2020.

16. Muniyappa R, Gubbi S. COVID-19 pandemic, coronaviruses, and diabetes mellitus. American journal of physiology Endocrinology and metabolism. 2020;318(5):E736–e41.

17. Barron E, Bakhai C, Kar P, Weaver A, Bradley D, Ismail H, et al. Associations of type 1 and type 2 diabetes with COVID-19-related mortality in England: a whole-population study. The lancet Diabetes & endocrinology. 2020;8(10):813–22.

18. Holman N, Knighton P, Kar P, O‘Keefe J, Curley M, Weaver A, et al. Risk factors for COVID-19-related mortality in people with type 1 and type 2 diabetes in England: a populationbased cohort study. The lancet Diabetes & endocrinology. 2020;8(10):823–33.

19. Navaratnam AV, Gray WK, Day J, Wendon J, Briggs TWR. Patient factors and temporal trends associated with COVID-19 in-hospital mortality in England: an observational study using administrative data. The Lancet Respiratory medicine. 2021;9(4):397–406.

20. Helms J, Kremer S, Merdji H, Clere-Jehl R, Schenck M, Kummerlen C, et al. Neurologic Features in Severe SARS-CoV-2 Infection. The New England journal of medicine. 2020.

21. Weiland NL S. Zimmer, C. U.S. calls for pause on Johnson & Johnson vaccine after clotting cases. New York Times. 2021 13 April 2021.

22. Haendel MA, Chute CG, Bennett TD, Eichmann DA, Guinney J, Kibbe WA, et al. The National COVID Cohort Collaborative (N3C): Rationale, design, infrastructure, and deployment. Journal of the American Medical Informatics Association : JAMIA. 2021;28(3):427–43.

